# Clinical and laboratory characteristics of clozapine treated schizophrenia patients referred to a national immunodeficiency clinic reveals a B-cell signature resembling CVID

**DOI:** 10.1101/19007815

**Authors:** M.J. Ponsford, R. Steven, K. Bramhall, M Burgess, S Wijetilleka, E. Carne, F McGuire, C. Price, M. Moody, S Zouwail, T Tahir, D. Farewell, T. El-Shanawany, S. Jolles

## Abstract

**Purpose:** An association between antibody deficiency and clozapine use in individuals with Schizophrenia has recently been reported. We hypothesized that if clozapine-associated hypogammaglobulinaemia was clinically relevant this would manifest in referral patterns.

**Methods:** Retrospective case note review of patients referred and assessed by Immunology Centre for Wales (ICW) between January 2005 and July 2018 with extraction of clinical and immunologic features for individuals with diagnosis of schizophrenia-like illness.

**Results:** 1791 adult patients were assessed at ICW during this period; 23 patients had a psychiatric diagnosis of schizophrenia or schizo-affective disorder. Principal indications for referral were findings of low calculated globulin and immunoglobulins. Clozapine was the single most commonly prescribed antipsychotic (17/23), disproportionately increased relative to reported use in the general schizophrenia population (OR 6.48, 95% CI: 1.79 to 23.5). Clozapine therapy was noted in 6/7 (86%) of patients subsequently requiring immunoglobulin replacement therapy (IgRT). Marked reduction of class-switched memory B-cells (CSMB) and plasmablasts were observed in clozapine-treated individuals relative to healthy age-matched controls. Clozapine duration is associated with CSMB decline. One patient discontinued clozapine, with gradual recovery of IgG levels without use of IgRT.

**Conclusion:** Our findings are consistent with enrichment of clozapine-treatment within schizophrenic individuals referred for ICW assessment over the last 13 years. These individuals displayed clinical patterns closely resembling the primary immunodeficiency CVID, however appears reversible upon drug cessation. This has diagnostic, monitoring and treatment implications for psychiatry and immunology teams and directs prospective studies to address causality and the wider implications for this patient group.

## Introduction

Schizophrenia is an enduring major psychiatric disorder affecting around 1% of the population (1). In addition to the debilitating psychiatric symptoms, it has major psychosocial consequences with an unemployment rate of 80-90% and a life expectancy reduced by 10-20 years (1) including suicide rates of approximately 5% (2). Societal costs in England alone are estimated to be £11·8 billion per year (3). Clozapine is a dibenzodiazepine atypical antipsychotic and the only licensed medication, for approximately 30% of patients with treatment-resistant schizophrenia (TRS) (1, 4). There is increasing evidence linking clozapine with pneumonia-related admissions (5-7) and mortality (8-11). Postulated mechanisms include sialorrhoea, sedation, agranulocytosis, and aspiration. We recently reported an association between clozapine therapy in schizophrenia and hypogammaglobulinemia (12, 13), greater than that reported following rituximab and methotrexate therapy in rheumatoid arthritis (14). To better define the clinical and immunologic abnormalities associated with clozapine use we performed a retrospective case review of patients assessed at the Immunology Centre for Wales (ICW).

## Methods

Electronic case records for patients assessed at ICW between January 2005 and July 2018 to identify all individuals with a concomitant psychiatric diagnosis of schizophrenia or schizo-affective disorder. Indication for referral, medication, and co-morbidities, and immunological testing at initial assessment and treatments were extracted using a standardised proforma. Recurrent infection history was defined as ≥3 distinct antibiotic courses per year or serious infection requiring admission, as in the wider literature (15).

All testing was performed in the United Kingdom Accreditation Service (UKAS) accredited Medical Biochemistry & Immunology Laboratory at the University Hospital of Wales. Immunoglobulin levels (IgG, IgA and IgM) were assayed by nephelometry (Siemens BN2 Nephelometer; Siemens), serum electrophoresis (Sebia Capillarys 2; Sebia, Norcross, GA, USA) and, where appropriate, serum immunofixation performed (Sebia Hydrasys; Sebia, Norcross, GA, USA). Antibody titres against haemophilus influenzae, tetanus and pneumococcal capsular polysaccharide were determined by enzyme-linked immunosorbent assay (The Binding Site, Birmingham, UK). Flow cytometry was performed using Beckman Coulter FC500 analyser. Lymphocyte phenotypes were analysed using Beckman Coulter Cyto-stat Tetrachrome reagents (CD45-FITC/CD4-RD1/CD8-ECD/CD3-PC5 and CD45-FITC/CD56-RD1/CD10-ECD/CD3-PC5), Flow-Count Fluorospheres and versalyse lysis solution. B cell phenotyping was performed as previously described (16) using the following antibodies: CD19-PE/Cy7 (Beckman Coulter), CD27-FITC (Serotec), CD21-PE (BD Pharmingen), CD38-FITC (Beckman Coulter), IgM Alexa-Flour 647 (Jackson ImmunoResearch), IgD-PE (Southern Biotech). CVID and age-matched healthy controls were analysed as part of an anonymous sample exchange scheme run jointly with King’s College London. Reference ranges are provided within the text. Individual clinical, immune, and treatment data are available online-**Supplementary, S1**.

### Statistical Analysis

Data was curated in Microsoft Excel. Fisher’s exact test and non-parametric Mann–Whitney U-testing, following D’Agostino & Pearson normality assessment, were conducted using GraphPad Prism v6.07. Where immunoglobulin level was undetectable, the lower limit of detection (IgG 1.34 g/L; IgA 0.05 g/L; and IgM 0.05 g/L) was used for data analysis, with density estimation and plotting performed in R (version 3.4.0). A two-tailed significance level of p<0.05 was used. This work was deemed as service evaluation by the institutional review board and requirement for ethical approval waived.

## Results

### Enrichment of clozapine-treated patients within schizophrenia cases referred for immunology assessment and requiring immunoglobulin replacement therapy

During the evaluation period 1791 adults were assessed at ICW; 23 had a diagnosis of schizophrenia or schizo-affective disorders. We hypothesized that if clozapine-associated hypogammaglobulinaemia was clinically relevant this would manifest in referral patterns. The mean clozapine prescription rate reported by the 2014 UK National Audit of Schizophrenia was 30% (17). We therefore expected a ratio of 7 Clozapine: 16 non-clozapine users. In contrast, we observed 17 patients with a history of clozapine use, corresponding to an odds ratio (OR) of 6.48 (95% CI: 1.79 to 23.5), p = 0.0072. This remained significant for prevalence estimates of clozapine use among the Welsh schizophrenia population up to 43% (**Supplementary, S2**). Patients receiving clozapine accounted for 6/7 (86%) of schizophrenia cases requiring IgRT, approximately 3% of our adult IgRT cohort. This compares to the international schizophrenia prevalence of 0.4-1% (18, 19). This suggests enrichment of clozapine-treated patients within our immunodeficiency cohort relative to the general population.

We next explored indication for referral and immunological finding at first assessment (individual patient details summarised-**Supplementary, S1**). Two clozapine-treatment patient with hypogammaglobulinaemia have been previously identified and reported (12). Recurrent infection was documented in 10/17 subjects (59%), predominately reflecting sinopulmonary infections. Four patients (24%) were referred with serum antibody levels below the 5^th^ percentile without any antibiotic use in the preceding 12 months (summarised-**Table 1**). A low calculated globulin (CG) (<23g/L) was present in 15/17 (88%) of subjects receiving clozapine and was associated with reductions in IgG, IgA, and IgM below the 5th percentiles in 14/17 subjects. We also found referral rates increased following the national introduction of CG screening to Wales during 2014, and prior to release of our initial report (12) (**Supplementary, S3**). Consistent with CG reduction, serum immunoglobulin levels were also reduced – summarised in **Figure 1**. Taken together, this is consistent with a clozapine-specific association with dysgammaglobulinaemia, and the utility of calculated globulin screening to stratify patients for specialist immunology assessment.

**Table 1:**
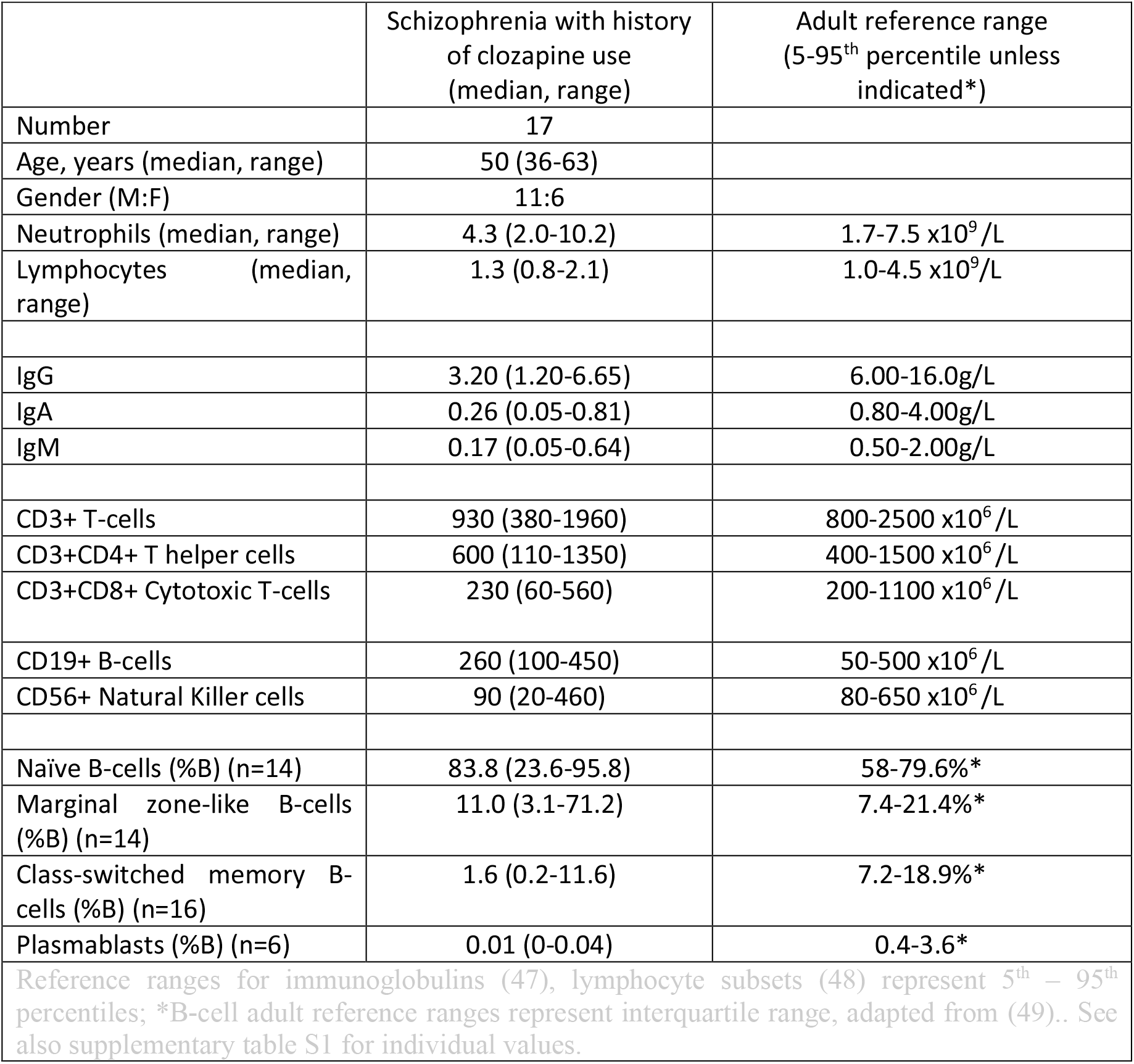
Immunology assessment summary at initial clinical visit. See supplementary S1 for individual patient values.

|  | Schizophrenia with history of clozapine use (median, range) | Adult reference range (5-95<sup>th</sup> percentile unless indicated*) |
| --- | --- | --- |
| Number | 17 |  |
| Age, years (median, range) | 50 (36-63) |  |
| Gender (M:F) | 11:6 |  |
| Neutrophils (median, range) | 4.3 (2.0-10.2) | 1.7-7.5 x10<sup>9</sup> /L |
| Lymphocytes (median, range) | 1.3 (0.8-2.1) | 1.0-4.5 x10<sup>9</sup>/L |
| IgG | 3.20 (1.20-6.65) | 6.00-16.0g/L |
| IgA | 0.26 (0.05-0.81) | 0.80-4.00g/L |
| IgM | 0.17 (0.05-0.64) | 0.50-2.00g/L |
| CD3+ T-cells | 930 (380-1960) | 800-2500 x10<sup>6</sup> /L |
| CD3+CD4+ T helper cells | 600 (110-1350) | 400-1500 x10<sup>6</sup> /L |
| CD3+CD8+ Cytotoxic T-cells | 230 (60-560) | 200-1100 x10<sup>6</sup> /L |
| CD19+ B-cells | 260 (100-450) | 50-500 x10<sup>6</sup> /L |
| CD56+ Natural Killer cells | 90 (20-460) | 80-650 x10<sup>6</sup> /L |
| Naïve B-cells (%B) (n=14) | 83.8 (23.6-95.8) | 58-79.6%* |
| Marginal zone-like B-cells (%B) (n=14) | 11.0 (3.1-71.2) | 7.4-21.4%* |
| Class-switched memory B-cells (%B) (n=16) | 1.6 (0.2-11.6) | 7.2-18.9%* |
| Plasmablasts (%B) (n=6) | 0.01 (0-0.04) | 0.4-3.6* |
| Reference ranges for immunoglobulins (47), lymphocyte subsets (48) represent 5<sup>th</sup> – 95<sup>th</sup> percentiles; *B-cell adult reference ranges represent interquartile range, adapted from (49).. See also supplementary table S1 for individual values. |  |  |

**Figure 1:**
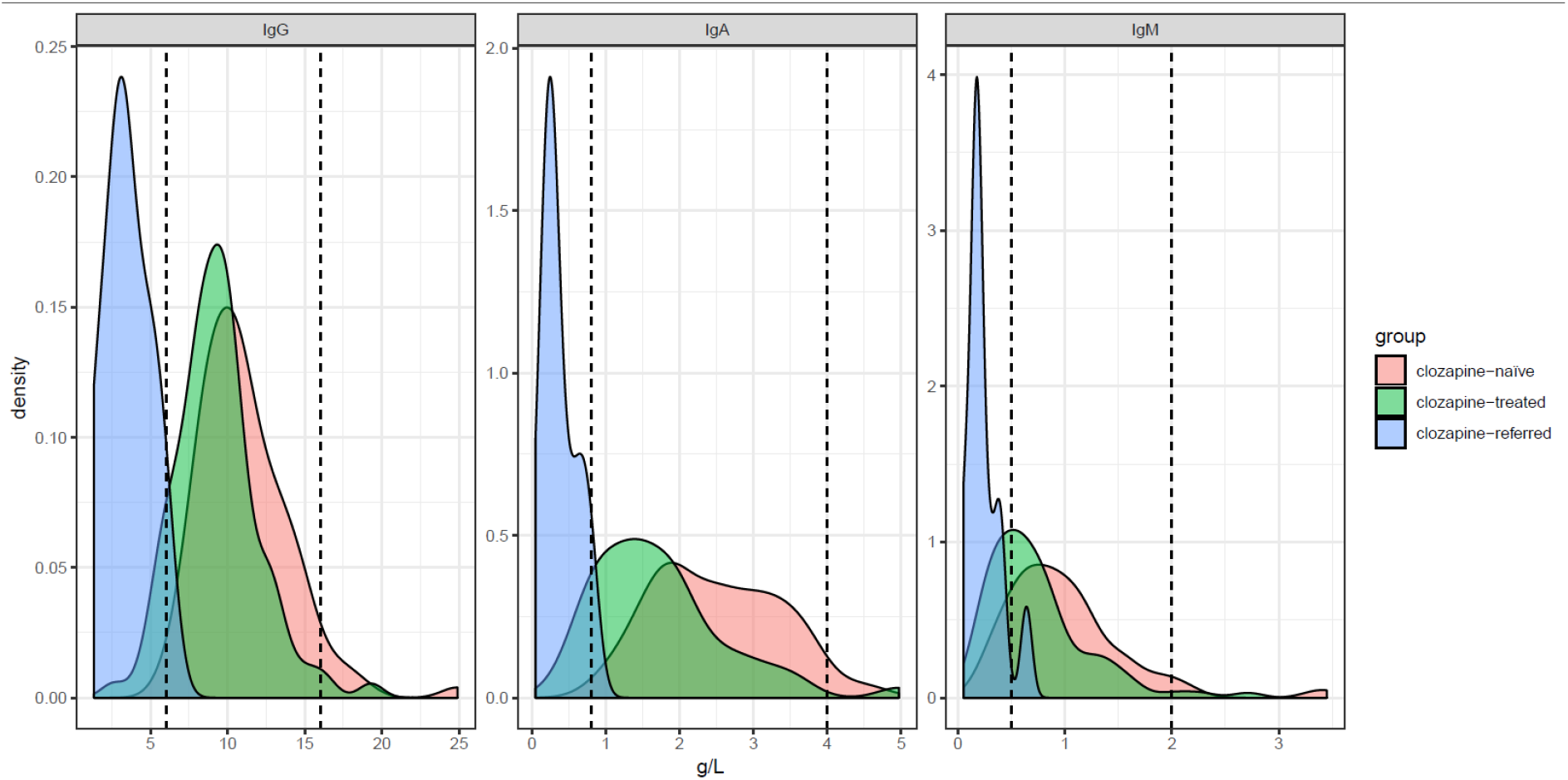
Immunoglobulin distribution in referred relative to referral cohort populations. Density plot showing distribution of serum immunoglobulin levels in patients receiving clozapine referred for Immunology assessment (light blue, n = 13 following removal of 4 patients n = 2 due to haematological malignancy and n = 2 previously included within the case-control study). Serum immunoglobulin distributions for clozapine-treated (green, n = 94) and clozapine-naive (pink, n = 98) antipsychotic treated Welsh schizophrenia cohorts are also shown for reference; adapted from (12). Dotted lines indicate the 5^th^ and 95^th^ percentiles for healthy adults.

*Comparison is made relative to the normal laboratory range (dotted lines, indicating 5-95*^*th*^ *percentiles) with comparison to immunoglobulin distributions in disease-control patient populations receiving clozapine or alternative antipsychotic agents and following exclusion of individuals with alternative causes of hypogammaglobulinaemia-from (12)*.

### B-cell dysregulation associated with long-term clozapine use resembles CVID

Clozapine-users are closely monitored for side effects of neutropenia and agranulocytosis, and all individuals showed normal neutrophil counts at first assessment. Lymphopenia and reduction in T-cells were present but largely confined to 2 individuals subsequently diagnosed with haematological malignancies **(Table 1; S2)**. Because the mechanisms underlying clozapine-associated hypogammaglobulinaemia remain unknown, we focused on B-cell clinical immunophenotyping data performed in isolation or as part of a full EUROClass evaluation (16). We took advantage of existing laboratory data to identify age-matched individuals with CVID (n=26) and healthy controls (n=16) to clozapine-treated patients (following exclusion of cases with haematological malignancy, n=15). Clozapine-treated individuals showed normal total B-cell counts but reduction of class-switched memory B-cells (CSMB) relative to healthy individuals (Mann-Whitney p< 0.0001), with the exception of a single patient who had discontinued clozapine after a 2-month trial and had normal immunoglobulin levels (**Figure 2; Supplementary S2**). We observed a general decline in CSMB with increasing duration of clozapine therapy. Immunophenotyping of plasmablasts was available for 6 individuals receiving clozapine and was comparable to CVID but significantly reduced compared to age-matched healthy controls (Mann-Whitney, p=0.004).

**Figure 2:**
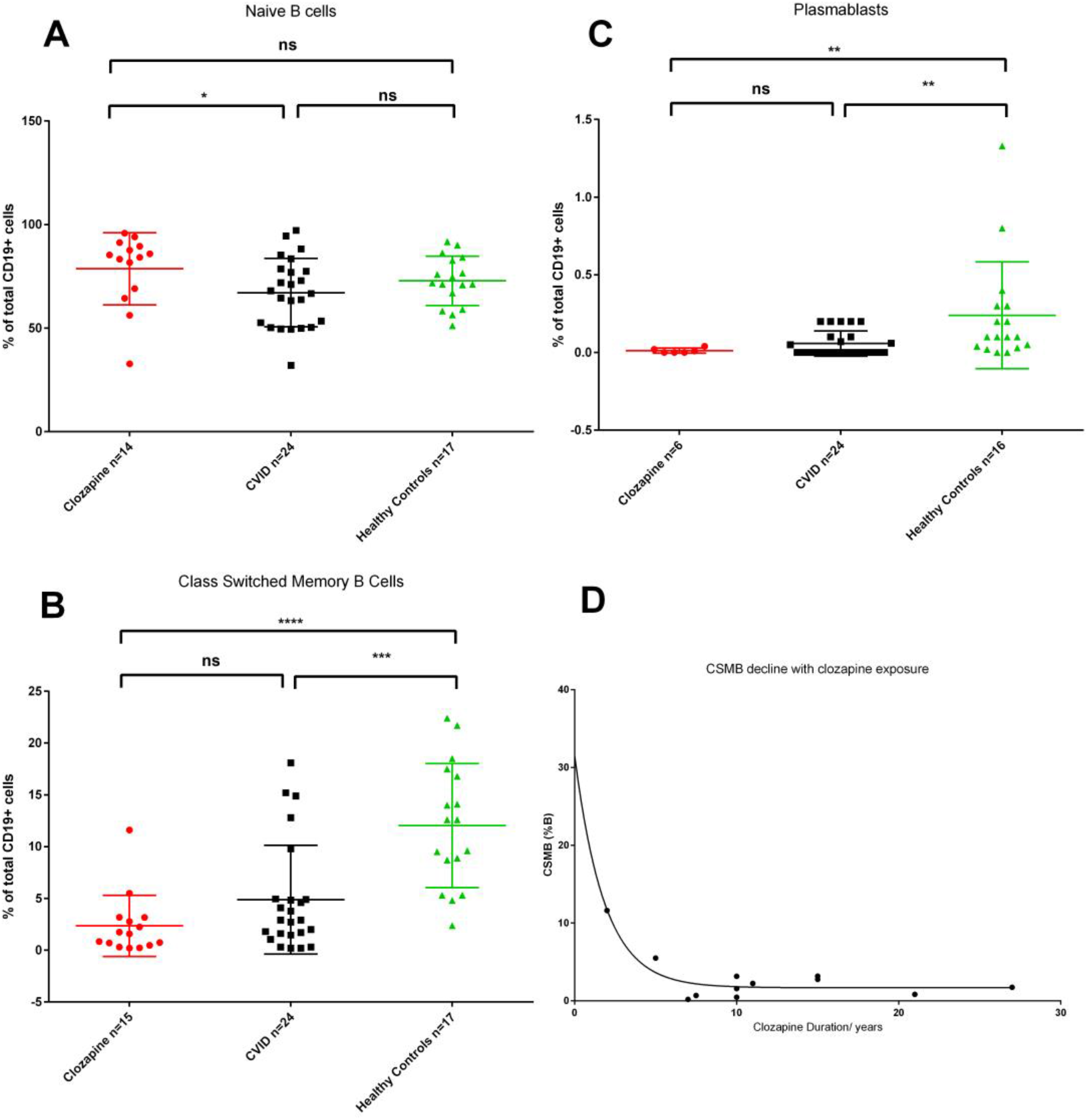
B-cell immunophenotyping in clozapine-treated patients and healthy controls. B cell subsets in patients with schizophrenia with history of clozapine therapy (numbers as shown), common variable immunodeficiency (CVID) and healthy controls. B-cell subsets gated on CD19+ cells and defined as follows: **A**: Naïve B-cells (CD27−IgD+IgM+), **B:** Class-switched Memory B-cells (CD27+IgD-IgM−), and **C:** Plasmablasts (CD21intCD38hiIgM+/-). Non-parametric Mann-Whitney testing performed given non-normally distributed data, *-p<0.05, **-p<0.01, ***-p<0.001, ****-p<0.0001; see text for probability values. **D**: Decline in CSMB levels relative to duration of clozapine exposure where available (data from 14 patients following exclusion of haematological malignancy cases where duration unclear) Line of best fit shown follows exponential decay modelling, R square = 0.8316.

Vaccine specific-IgG responses are routinely evaluated as part of clinical assessment (20). At initial assessment, levels below putative protective thresholds were common with IgG to Haemophilus influenza B (HiB) < 1mcg/ml (21) in 12/16 assessed (75%); Pneumococcus-IgG < 50mg/L (22) in 15/16 (94%); and Tetanus-IgG < 0.1 IU/mL in 6/16 (38%) individuals tested. Post-*Menitorix* (HiB/MenC conjugated to tetanus toxoid) vaccination serology was assessed after 4 weeks, with 5/11 (45%) individuals failing to mount a Haemophilus-IgG response ≥1mcg/ml, and 1/12 failing to exceed the ≥0.10 IU/mL post-vaccination Tetanus-IgG level defined by the World Health Organisation (23). Following *Pneumovax II*, 6/16 (38%) individuals failed to develop an IgG response above a threshold of ≥50mg/L (22).

***Supplementary, S1 and S4: vaccine response assessment***

### Reduction in infection burden following immunoglobulin replacement therapy

Our approach to antibody deficiency follows that previously reported (15, 20). Cumulative follow-up for clozapine-associated hypogammaglobulinaemia now exceeds 61 patient-years (range 0.5-11 years), during which time 6/16 (38%) of clozapine-treated patients were commenced on IgRT. Over a 12-month period prior to replacement, these patients received a mean of 6.5 acute antibiotic courses (range 3-12, including 1 inpatient admission requiring parenteral therapy). Following IgRT, this fell to a median of 1.5 oral antibiotics courses per year (range 0-2 courses), with no infection-triggered hospital admissions. Replacement trough IgG levels ranged from 5.5 to 9.6g/L (mean 8.0g/L).

### Partial recovery of immunoglobulin following clozapine discontinuation

One patient (supplementary table S1: IDs#7) discontinued clozapine due to the known side-effect of neutropenia, detected by the clozapine monitoring program, providing a unique opportunity to examine reversibility of humoral dysregulation. In the absence of IgRT Patient #7 has demonstrated a gradual recovery in terms of serum IgG level from 3.5g/L to 5.95g/L over 3 years, however IgA and IgM levels have remained depressed (**Figure 3**). Patient #2 discontinued clozapine 2 years ago, having already commenced IgRT, with subsequent recovery of IgM from 0.22 to 0.86. IgA levels have remained below 0.24g/L. Together this provides support for reversibility in humoral function following clozapine-withdrawal, but suggests this process is gradual and limited. In contrast, clozapine cessation in patient #2 was associated with profound and acute relapse of psychiatric symptoms, requiring prolonged inpatient admission to a specialist mental health unit.

**Figure 3:**
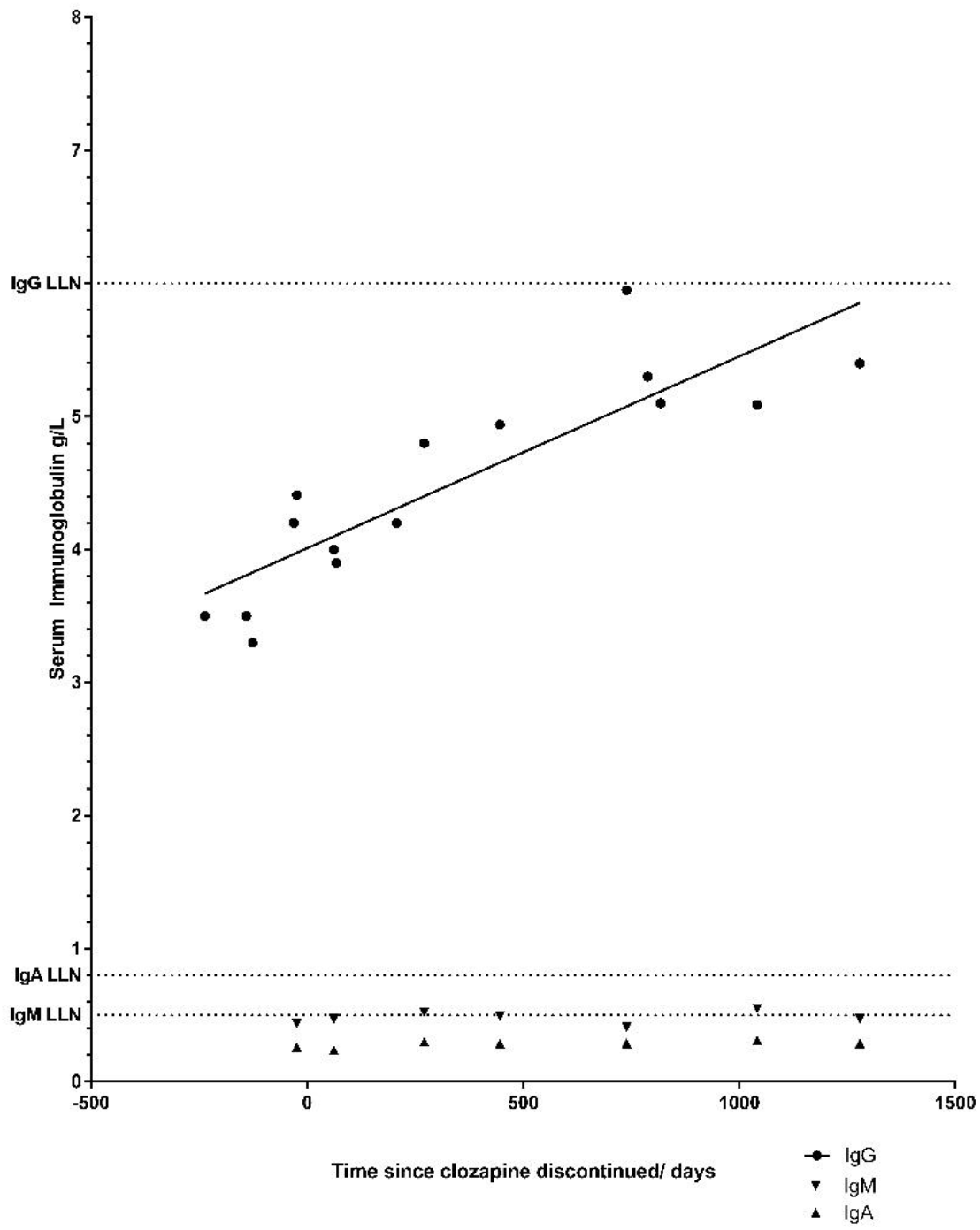
Gradual recovery of serum IgG post-discontinuation of Clozapine in a single patient without recourse to immunoglobulin replacement. LLN= lower limit of normal.

**Figure 3**: Gradual recovery of serum IgG post-discontinuation of clozapine in absence of immunoglobulin replacement.

## Discussion

In this retrospective review of referrals to a national immunology centre, we found that individuals with schizophrenia receiving clozapine displayed clinically significant panhypogammaglobulinemia, impaired vaccine responses and disturbed B cell maturation with a proportion benefitting from IgRT. Consistent with a specific relationship to clozapine-use, we observed enrichment of clozapine-use amongst schizophrenia patients undergoing immunology assessment or requiring IgRT relative to that reported in the wider population by a number of recent studies (4, 17, 24). The most common feature at referral was a low calculated globulin (25). Importantly, both clozapine and non-clozapine antipsychotic medications require routine liver function monitoring (26), arguing against a referral-bias due to differential testing. While the use of calculated globulin screening is being developed in a number of countries, a lack of wider availability may limit the identification of antibody deficient patients taking clozapine in other settings (25, 27, 28).

Our finding of a temporal association between clozapine use and reduction in CSMB is consistent with our previous analysis of a separate cohort that hypogammaglobulinaemia appears a late manifestation of clozapine therapy (12). Our immunological findings associated with clozapine-associated hypogammaglobulinaemia is independently supported by a recently reported peripheral blood immune signature for TRS patients revealing reduced plasmablasts and CSMB relative to disease and healthy controls (29). Whilst this study focused on psychiatric status, the majority of such patients were clozapine-treated; immunoglobulin levels were not determined by this group (29). These findings also suggest that many clozapine-treated patients in our cohort and beyond may have received a diagnosis of CVID, according to internationally-accepted criteria (30). We are conducting a wider survey of UK Immunology centres to examine this observation.

This study has several limitations, including its sample size and single-centre retrospective design. Prospective studies are required, for instance to validate the association between therapy duration and effect on B-cell phenotypes, and to address causality. We speculate an effect targeting late stage B-cell differentiation or survival may underlie the observed findings (31). However, at present, we cannot exclude a link to the underlying pathophysiology of schizophrenia-which like CVID remains largely unknown. Intriguingly, pathway analysis of schizophrenia associations have demonstrated marked enrichment at enhancers active in immune tissues, particularly B-lymphocyte lineages (32). Polypharmacy, multi-morbidity, and socio-economic status are common and important factors complicating healthcare delivery in schizophrenia (33, 34) and might also contribute to the observed findings. Smoking is common in both clozapine-treated and clozapine-naïve schizophrenia patients (35), predisposes to pneumonia (36), and may lead to systemic steroid therapy for chronic obstructive pulmonary disorder. However, the reduced CSMB and pan-hypogammaglobulinaemia profiles we observed clearly differ from the patterns previously reported with both steroids or smoking (37, 38). Other medication including metformin and anti-epileptics have been described to have immunomodulatory effects in murine studies (39) or case series (40) however do not fully explain the observed hypogammaglobulinaemia -in line with the rigorous exclusion criteria applied during our initial case-control study (12). The immunomodulatory effects of the dibenzodiazepine clozapine are increasingly recognised beyond agranulocytosis and neutropenia (41-43). Here, we present evidence of gradual recovery of humoral immune function following clozapine withdrawal in the absence of IgRT.

From a practical standpoint, recovery of immunoglobulin levels appeared gradual and variable. Weighed against the florid relapse in psychiatric symptoms observed following clozapine discontinuation in one, we do not currently suggest discontinuation of clozapine in patients. This is in line with the unique efficacy of clozapine in TRS (44), and availability of treatment options to mitigate the risks associated with antibody deficiency. Finally, given evidence of increased risk of pneumonia within individuals with schizophrenia (7, 45, 46) our finding of low levels of baseline immunity to common vaccinations within this patient group highlights a simple strategy for risk mitigation.

### Summary

We report the clinical features of clozapine-associated immunodeficiency identified following the introduction of calculated globulin screening in Wales and highlight clinically significant panhypogammaglobulinemia, impaired vaccine responses, and new findings of disturbed B-cell maturation. In support of a drug-related effect we show a temporal association between clozapine exposure and reduction in CSMB levels, and evidence of gradual reversibility following discontinuation. Clinicians should be alert for this diagnostic overlap with CVID. Patients may benefit from monitoring or clinical intervention in the form of vaccination, antibiotic prophylaxis, or immunoglobulin replacement therapy (IgRT). Clozapine’s immunomodulatory effects are poorly understood, and further studies are required to delineate mechanism.

## Data Availability

Supplementary information also uploaded

## Highlights Box

- **What is known about this topic already**
- Clozapine remains the only effective medication for treatment-resistant schizophrenia (TRS) however is associated with an increased risk of pneumonia and death. Clozapine associated antibody-deficiency has only recently been reported in this patient group.
- **What does this article add to our knowledge**
- Hypogammaglobulinaemia associated with TRS-patients receiving clozapine can result in a serious and significant infection burden which in Immunodeficiency Clinic assessment was associated with impaired vaccine responses, low class-switched memory B-cells and plasmablasts resembling primary immunodeficiency. Patients often required intervention in the form of antibiotic prophylaxis and/or immunoglobulin replacement (IgRT).
- **How does this study impact current management guidelines**
- Clinicians need to be aware of the association between clozapine and antibody deficiency. Such patients may currently be diagnosed with CVID.
- Current clozapine monitoring does not include antibody testing however calculated globulin may be a helpful screening tool.
- A number of potential interventions may help risk mitigate clozapine-associated antibody deficiency.

## Disclosures

MP is supported by a Welsh Clinical Academic Training Fellowship and Wellcome Trust Institutional Strategic Support Fund (ISSF) grant.

SJ has received support from CSL Behring, Shire, LFB, Biotest, Binding Site, Sanofi, GSK, UCB Pharma, Grifols, BPL SOBI, Weatherden, Zarodex and Octapharma for projects, advisory boards, meetings, studies, speaker, and clinical trials.

TE has received educational support, project support, advisory board fees, speaker fees and/or clinical trial support from Biotest, CSL, LFB, Mylan, Novartis, Shire and Werfen.

CP has received support to attend educational meetings from CSL Behring and ALK.

The remaining authors have no relevant conflicts of interest to declare.

## Acknowledgements

We gratefully acknowledge the patients and wider clinical immunology and mental health teams who have contributed, and Dr Mohammad Ibrahim and the King’s College Hospital Immunology team for participating in immunophenotyping sample exchange schemes.

## Abbreviations

CSMB: Class switched memory B-cells
CVID: Common variable immunodeficiency
EAE: Experimental autoimmune encephalitis
IgRT: Immunoglobulin replacement therapy
22q11ds: 22q11 deletion syndrome (also known as DiGeorge)
TRS: Treatment resistant schizophrenia

---

**Supplementary S1: Clinical and Immunological Information for individual clozapine-treated patients**

- Attached as separate .xls file

**Supplementary S2: Contingency Table Outcomes modelling difference prevalence of clozapine use within the Welsh Schizophrenia population**

- Attached as separate .docx file

**Supplementary Figure, S3:**
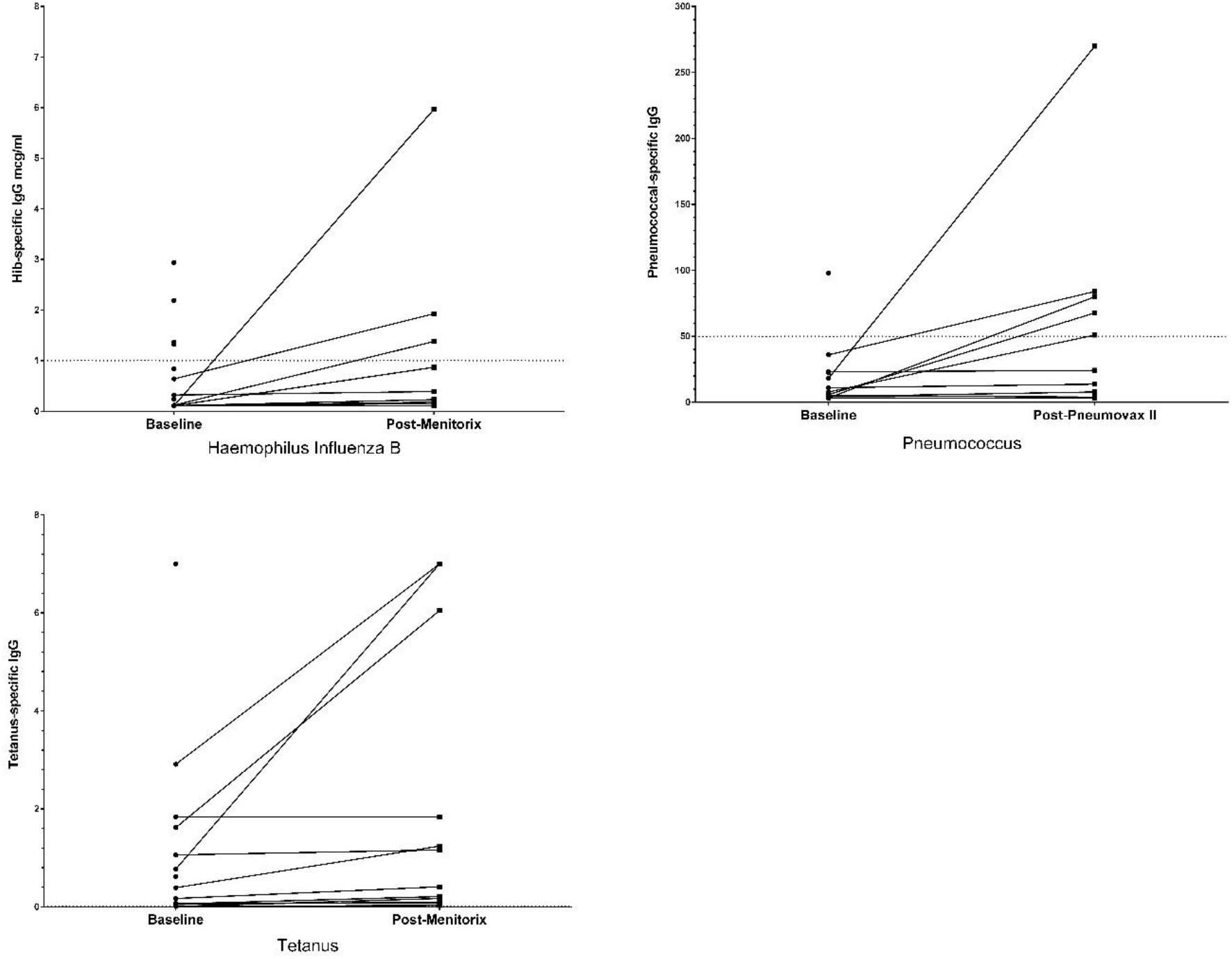
Vaccine specific-IgG response assessment. See text and supplementary S1 for details.

**Supplementary, S4:**
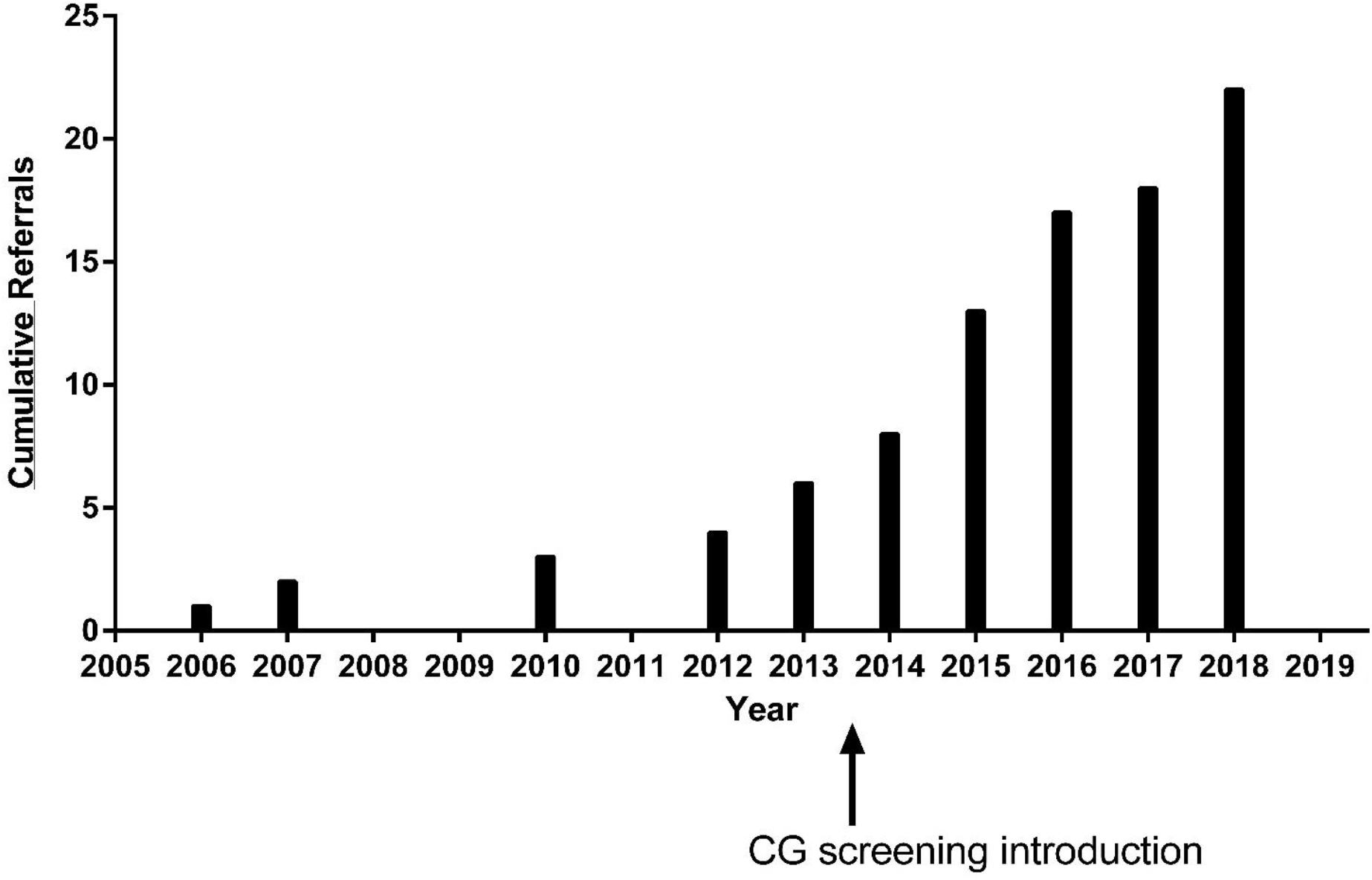
Cumulative Referrals to ICW with a diagnosis of schizophrenia in relation to introduction of calculated globulin screening. The national calculated globulin screening programme was setup during 2014, with roll-out complete across all health boards by November 2014. Consequently, any adult liver function test with low calculated globulin triggers the laboratory comment “*Low calculated globulin-may represent antibody deficiency. Consider immunoglobulins if there is a history of infections*”.

## Kudos Plain Language Summary

The introduction of clozapine in the 1950s was a major therapeutic advance in the treatment of schizophrenia. It remains the gold standard therapy for approximately 3/10 individuals who fail to respond to initial management. Overall clozapine improves symptoms and saves lives. Recent studies (2018) have suggested that clozapine therapy may be associated with a block in antibody production (causing antibody-deficiency). This would predict patients receiving this therapy to be more likely to experience infections (such as pneumonia). This work examines patients referred to a major immunology centre since 2005, with focus on those receiving antipsychotic medicines (including clozapine). As predicted by the authors (but against the normal pattern of prescribing), clozapine was the most common antipsychotic used by patients referred with antibody deficiency. The authors go on to define the clinical and immunological features of this group, highlighting a close similarity to individuals without a known cause of antibody deficiency. Several clozapine-treated patients went on to receive antibody replacement therapy, successfully reducing their infection rate. By following these patients over time, they also saw that in one patient who stopped clozapine treatment that this was associated with a gradual return in their major antibody (IgG). This adds support to the argument that clozapine therapy is associated with a drug-related (secondary) antibody deficiency. This is an important finding for immunologists around the work, who could easily confuse a drug-related cause of antibody deficiency with an inherited (primary) case. The authors remain cautious and suggest further studies are needed with greater size and following patients before and after they start/stop clozapine. This is because the disease processes underlying both schizophrenia and primary antibody deficiency remain largely unknown: meaning there could be important shared mechanisms linking both conditions.

## Notes

### Author Declarations

All relevant ethical guidelines have been followed and any necessary IRB and/or ethics committee approvals have been obtained.

Any clinical trials involved have been registered with an ICMJE-approved registry such as ClinicalTrials.gov and the trial ID is included in the manuscript.

